# Genetic susceptibility to earlier ovarian ageing increases *de novo* mutation rate in offspring

**DOI:** 10.1101/2022.06.23.22276698

**Authors:** Stasa Stankovic, Saleh Shekari, Qin Qin Huang, Eugene J. Gardner, Nick D. L. Owens, Ajuna Azad, Gareth Hawkes, Katherine A. Kentistou, Robin N. Beaumont, Felix R. Day, Yajie Zhao, The Genomics England Research Consortium, Kitale Kennedy, Andrew R. Wood, Michael N. Weedon, Ken K. Ong, Caroline F. Wright, Eva R. Hoffmann, Matthew E. Hurles, Katherine S. Ruth, Hilary C. Martin, John R. B. Perry, Anna Murray

## Abstract

Human genetic studies have provided substantial insight into the biological mechanisms governing ovarian ageing, yet previous approaches have been largely restricted to assessing common genetic variation. Here we report analyses of rare (MAF<0.1%) protein-coding variants in the exomes of 106,973 women from the UK Biobank study, implicating novel genes with effect sizes up to ∼5 times larger than previously discovered in analyses of common variants. These include protein truncating variants in *ZNF518A*, which shorten reproductive lifespan by promoting both earlier age at natural menopause (ANM, 5.61 years [4.04-7.18], *P*=2*10^-12^) and later puberty timing in girls (age at menarche, 0.56 years [0.15-0.97], *P*=9.2*10^-3^). By integrating ChIP-Seq data, we demonstrate that common variants associated with ANM and menarche are enriched in the binding sites of *ZNF518A*. We also identify further links between ovarian ageing and cancer susceptibility, highlighting damaging germline variants in *SAMHD1* that delay ANM and increase all-cause cancer risk in both males (OR=2.1 [1.7-2.6], *P*=4.7*10^-13^) and females (OR=1.61 [1.31-1.96], *P*=4*10^-6^). Finally, we demonstrate that genetic susceptibility to earlier ovarian ageing in women increases *de novo* mutation rate in their offspring. This provides direct evidence that female mutation rate is heritable and highlights an example of a mechanism for the maternal genome influencing child health.

## Introduction

Reproductive longevity in women varies substantially in the general population, and has a profound impact on fertility and health outcomes in later life^1–3^. Women are born with a non-renewable ovarian reserve, which is established during foetal development. This reserve is continuously depleted throughout reproductive life, ultimately leading to menopause^4–6^. Variation in menopause timing is largely dependent on the differences in the size of the initial oocyte pool and the rate of follicle loss^3^. Natural fertility is believed to be closely associated with menopause timing, and it declines on average 10 years before the onset of menopause^4,7^. The effect of early menopause on infertility is becoming increasingly relevant due to the secular trend of delaying parenthood to later maternal age at childbirth, especially in Western populations. In addition, normal variation in reproductive lifespan is causally associated with the risk of a wide range of disease outcomes, such as type 2 diabetes mellitus, cancer and impaired bone health, further highlighting the need for better understanding of the regulators and physiological mechanisms involved in reproductive ageing^1,8^.

The variation in timing of menopause reflects a complex mix of genetic and environmental factors that population-based studies have begun to unravel. Previous genome-wide association studies (GWAS) have successfully identified ∼300 distinct common genomic loci associated with the timing of menopause^1^. These reported variants cumulatively explain 10% - 12% of the variance in ANM and 31-38% of the overall estimated SNP heritability^1,9,10^. The majority of these loci implicate genes that regulate DNA damage response (DDR), highlighting the particular sensitivity of oocytes to DNA damage due to the prolonged state of cell cycle arrest across the life-course^1,7,11–20^.

Genetic studies for ANM to date have largely focussed on assessing common genetic variation, with little insight into the role of rarer, protein-coding variants. Initial exome-sequencing (WES) analyses in UK Biobank identified gene-based associations with ANM for *CHEK2, DCLRE1A, HELB, TOP3A, BRCA2* and *CLPB*^1,9^. In this study, we aimed to explore the role of rare damaging variants in ovarian ageing in greater detail through a combination of enhanced phenotype curation, better powered statistical tests and assessment of different types of variant classes at lower allele frequency thresholds (**Supplementary Note**). Using these approaches we identify five genes harbouring variants of large effect that have not previously been implicated, highlighting *ZNF518A* as a major transcriptional regulator of ovarian ageing. Furthermore, we extend these observations to show that women at increased genetic risk of earlier menopause have increased rates of *de novo* mutations in their offspring.

## Results

### Exome-wide gene burden associations with ANM

To assess the impact of rare damaging variants on age at natural menopause (ANM), we used whole-exome sequencing (WES) data available in 106,973 post-menopausal UK Biobank female participants of European genetic-ancestry^21^. Individual gene burden association tests were conducted by collapsing genetic variants according to their predicted functional categories. We defined three categories of rare exome variants with minor allele frequency (MAF) < 0.1%: high-confidence Protein Truncating Variants (HC-PTVs), missense variants with CADD score ≥ 25, and ‘damaging’ variants (defined as combination of HC-PTVs and missense variants with CADD ≥ 25). We analysed 17,475 protein-coding genes with the minimum of 10 rare allele carriers in at least one of the masks tested. The primary burden association analysis was conducted using BOLT-LMM^22^ (**Supplementary Table 1**). The low exome-wide inflation scores (e.g. PTV λ=1.047) and the absence of significant association with synonymous variant burden for any gene indicate our statistical tests are well calibrated (**Supplementary Figure 1**).

We identified rare variation in nine genes associated with ANM at exome-wide significance (*P*<1.08*10^-6^, **Figures 1 and 2, Supplementary Figures 2 and 3**). These were confirmed by an independent group of analysts using different QC and analysis pipelines (**Supplementary Tables 1, 2**). Three of these genes have been previously reported in UKBB WES analysis^9^ - we confirm the associations of *CHEK2* (beta=1.57 years, 95% CI: 1.23-1.92, *P*=1.60*10^-21^, N=578 damaging allele carriers) and *HELB* (beta=1.84, 95% CI: 1.08-2.60, *P*=4.20*10^-7^, N=120 HC-PTV carriers) with later ANM and a previously borderline association of *HROB* with earlier ANM (beta= -2.89 years, 95% CI: 1.86-3.92, *P*=1.90*10^-8^, N=65 HC-PTV carriers). In addition, our previous ANM GWAS analyses^1^ identified an individual low-frequency PTV variant in *BRCA2*, which we now extend to demonstrate that, in aggregate, *BRCA2* HC-PTV carriers exhibit 1.18 years earlier ANM (beta= -1.18, 95% CI: 0.72-1.65, *P*=2.60*10^-7^, N=323). Rare variants in the remaining five genes – *ETAA1, ZNF518A, PNPLA8, PALB2* and *SAMHD1* have not been previously implicated in ovarian ageing. Effect sizes of these associations range from 5.61 years earlier ANM for HC-PTV carriers in *ZNF518A*, to 1.35 years later ANM for women carrying damaging alleles in *SAMHD1*. This contrasts with a maximum effect size of 1.06 years (median 0.12 years) for common variants (MAF>1%) identified by previous ANM GWAS^1^.

**Figure 1:**
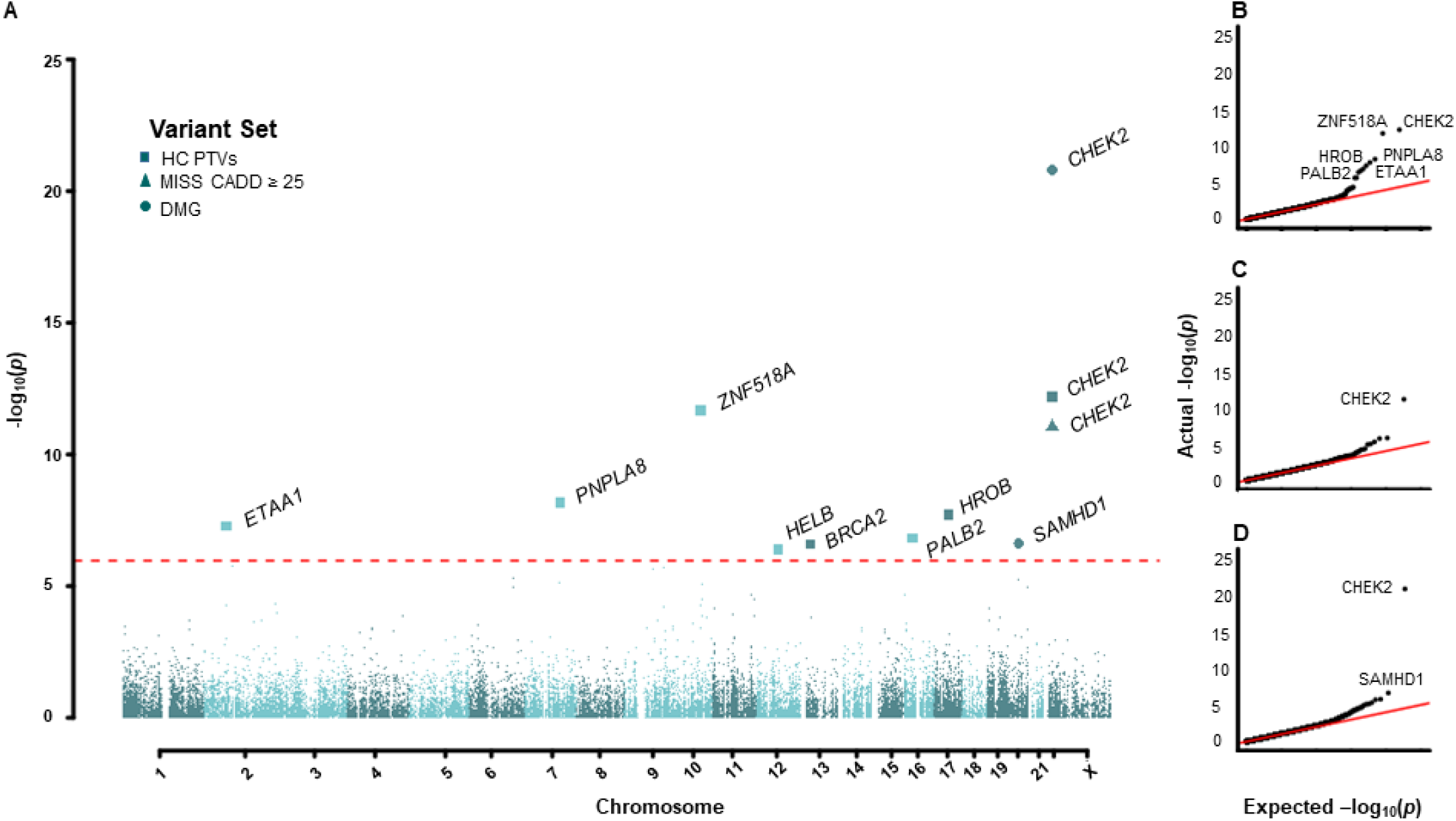
Exome-wide associations with age at natural menopause. **(A)** Manhattan plot showing gene burden test results for age at natural menopause. Genes passing exome-wide significance (P<1.08*10^-6^) are indicated, with point shape signifying the variant class tested. **(B-E)** QQ plots for **(B)** high confidence PTVs **(C)** CADD ≥ 25 missense variants **(D)** damaging variants.

**Figure 2:**
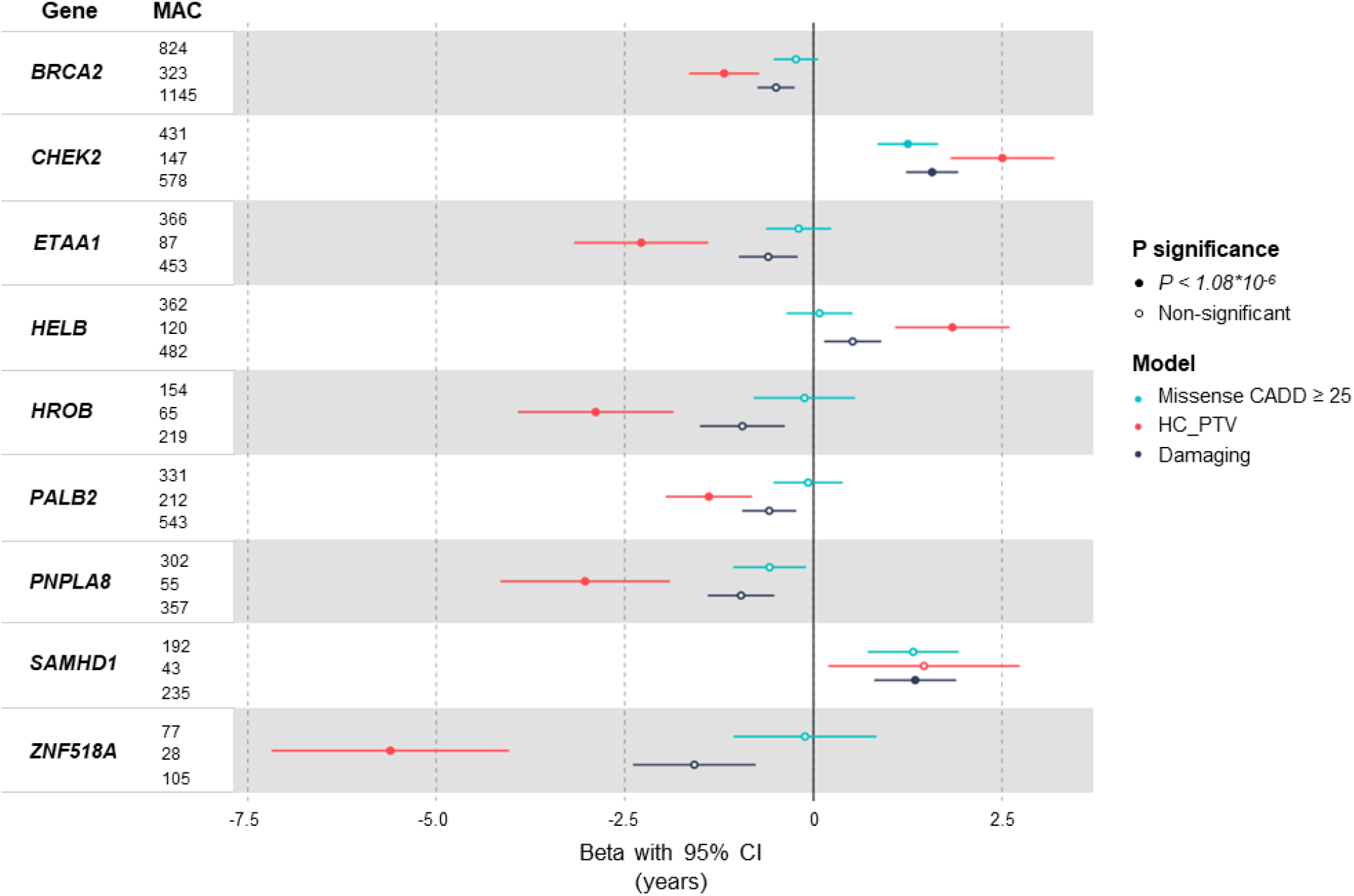
Forest plot for gene burden associations with age at natural menopause. Exome-wide significant (P < 1.08*10^-6^) genes are displayed. Points and error bars indicate beta and 95% CI for the variant category indicated. Betas, CIs, Minor Allele Counts (MAC) and P values are derived from BOLT-LMM.

We next sought to understand why previous analyses of UKBB WES data missed the associations we report here, and conversely why we did not identify associations with other previously reported genes. Of the seven genes identified by Ward *et al*.^9^, three were also identified by our study (*CHEK2, HELB* and *HROB*), three were recovered when we increased our burden test MAF threshold from 0.1% to 1% (*DCLRE1A, RAD54L, TOP3A*), and an additional gene fell just below our *P* value threshold when considering variants with <1% MAF (*CLPB; P =1*.*2*10*^-5^). In contrast, our discovery of novel associations that were not reported by Ward *et al*. (*BRCA2, ETAA1, PALB2, PNPLA8, SAMHD1* and *ZNF518A*) were likely explained by differences in phenotype preparation, sample size, variant annotation and the statistical model used (see **Supplementary Note and Supplementary Table 3**).

### Exploring common variant associations at identified ANM genes

To explore the overlap between common and rare variant association signals for ANM, we integrated our exome-wide results with data generated from the largest reported common variant GWAS of ANM^1^.

Five of our nine identified WES genes (*CHEK2, BRCA2, ETAA1, HELB* and *ZNF518A)* mapped within 500kb of a common GWAS signal (**Supplementary Table 4**). Notably, we previously reported a common, predicted benign, missense variant (rs35777125-G439R, MAF=11%) in *ETAA1* associated with 0.26 years earlier ANM. In contrast, our WES analysis identified that carriers of rare HC-PTVs in *ETAA1* show a nearly 10-fold earlier ANM (beta= -2.28 years, 95% CI: 1.39-3.17, *P*=5.30*10^-8^, N=87). Furthermore, three independent non-coding common GWAS signals ∼150kb apart (MAF: 2.8-47.5%, beta: -0.28-0.28 years per minor allele) were reported proximal to *ZNF518A*, whereas gene burden testing finds that rare HC-PTV carriers show nearly 20-fold earlier ANM than common variant carriers (beta= -5.61 years, 95% CI: 4.04-7.18, *P*=2.10*10^-12^, N=28).

In addition there were two genes within 500kb of GWAS loci (*BRCA1* and *SLCO4A1*) that were associated with ANM by gene burden testing at *P*<1.7*10^-5^. Effect sizes for common variant associations ranged from 0.07-0.24 years per allele at these loci, whereas gene burden tests for rarer variants at these same loci revealed much larger effect sizes: for *BRCA1*, 2.1 years earlier for PTVs (*P=2*.*4*10*^*-6*^) and for *SLCO4A1*, 1.13 years earlier ANM for damaging variants *(P=1*.*1*10*^*-5*^), with non-overlapping 95% confidence intervals between common and rare variant associations for *BRCA1*.

### Common ANM associated variants are enriched in *ZNF518A* binding sites

Heterozygous loss of function of *ZNF518A* had the largest effect on ANM of the genes we identified. *ZNF518A* is a poorly characterised C2H2 zinc finger transcription factor, which has been shown to associate with PRC2 and G9A-GLP repressive complexes along with its paralog *ZNF518B*, suggesting a potential role in transcriptional repression^23^. *ZNF518A* localises robustly to 18,706 sites in the genome, based on ChIP-seq data available from ENCODE^24,25^ and binds primarily to gene promoters, with 33.5% (6,263) of *ZNF518A* binding sites within 2kb of a transcription start site (TSS) (**Supplementary Figure 4a-c**). Common variants associated with ANM^1^ were enriched in the transcriptional targets of *ZNF518A* (*P*=1.32*10^-4^) using fGWAS^26^. We further tested functional enrichment using signed linkage disequilibrium profile (SLDP) regression^27^. This confirmed the enrichment of *ZNF518A* binding sites near to loci associated with ANM and showed that its transcriptional repression is associated with earlier ANM (*P*=0.02), consistent with evidence from rare variant burden tests. Separating *ZNF518A* sites by those proximal (< 2Kb) and distal (>5kb) from a TSS, demonstrated this association was due to *ZNF518A* binding at regulatory regions distal to the TSS (proximal TSS *P*=0.3, distal *ZNF518A P*=0.002). Notably, these regulatory *ZNF518A* bound loci produce the largest association amongst an SLDP catalogue of 382 transcription factors and regulators (**Supplementary Table 5, Supplementary Figure 4d**). These results suggest a different functional role for *ZNF518A* at TSS and more distal regulatory regions. In order to explore this further we assessed the sequence determinants of *ZNF518A* binding. *De novo* motif discovery identified an AT-rich motif enriched at distal regulatory *ZNF518A* binding sites, but not at TSS bound by *ZNF518A*. This AT-rich motif was centrally enriched within *ZNF518A* ChIP-seq peaks, and matched an unvalidated motif present in the JASPAR transcription factor motif database^28^ (**Supplementary Figure 4e**). We found the number of perfect instances of this AT-rich motif to be strongly associated with *ZNF518A* occupancy as assessed by *ZNF518A* ChIP-seq signal at distal regions but not at TSS (**Supplementary Figure 4f,g**). At distal regions, the maximal association between peaks greater than the median height was found at least seven motif instances (Hypergeometric right tail *P* < 10^-389^, Odds Ratio 7.41). These data suggest that *ZNF518A* is recruited by DNA sequence at distal sites, but at TSS may be recruited to gene promoters by interaction with another DNA binding factor.

We next employed public *in vitro* differentiated human primordial germ like-cell data^29,30^ to assess the chromatin state at *ZNF518A* bound loci, directly comparing distal regions with TSS. *ZNF518A* bound TSS showed chromatin accessibility^30^ and were marked with H3K27ac^29^. In contrast, distal regions lacked H3K27ac and showed minimal chromatin accessibility **(Supplementary Figure 4h)**. Extending this comparison to the Epimap chromatin states^31^, we find that overall *ZNF518A* bound loci are enriched in active TSS and that distal *ZNF518A* regions are variously enriched in active and repressed chromatin **(Supplementary Figure 4i,j)**. Consistent with previous data which has found *ZNF518A* in repressive complexes, these data suggest that *ZNF518A* is recruited by DNA sequence to distal regulatory regions where it acts to repress local chromatin.

While *ZNF518A* is known to have diverse tissue expression including the ovary, we found that it was highly expressed in fetal germ cells at both the mitotic and meiotic stages (**Supplementary Tables 6 and 7; Supplementary Figures 5 and 6**). The eight other WES genes identified in this study were expressed at varying levels in fetal gonadal cells, oocytes and granulosa cells across different developmental stages (**Supplementary Figures 5 and 6**).

### Identified genes influence other aspects of health and disease

Our genetic studies have previously shown that the genetic mechanisms regulating the end of reproductive life are largely distinct from those determining its beginning^32,33^. However, it is noteworthy that the largest reported GWAS for age at menarche identified a common variant signal at the *ZNF518A* locus for later puberty timing in girls (rs1172955, beta= 0.04 years, 95% CI: 0.03-0.05, *P*=6.6*10^-12^), which appears nominally associated with earlier ANM (beta=-0.04, 95% CI: 0.01-0.06, *P*=6.6*10^-3^)^32^. To extend this observation, we found that our identified *ZNF518A* PTVs were also associated with later age at menarche (0.56 years, 95% CI: 0.14-0.98, *P*=9.2*10^-3^). Furthermore, using fGWAS and SLDP, we discovered that, similar to ANM, common variants that influence puberty in girls were enriched in transcriptional targets of *ZNF518A* (**Supplementary Table 5**). These data suggest that loss of *ZNF518A* shortens reproductive lifespan, by delaying puberty and reducing age at menopause.

We next explored what impact ANM-associated genes had on cancer outcomes and found a novel association of *SAMHD1* damaging variants and HC-PTVs with ‘All cancer’ in both males (OR=2.12, 95% CI: 1.72-2.62, *P*=4.7*10^-13^) and females (OR=1.61, 95% CI: 1.31-1.96, *P*=4*10^-6^; **Figure 3, Supplementary Table 8-10**). In addition we replicated previously reported associations with protein truncating variants in *BRCA2, CHEK2* and *PALB2* and cancer outcomes in males and females (**Supplementary Tables 8-10**).

**Figure 3:**
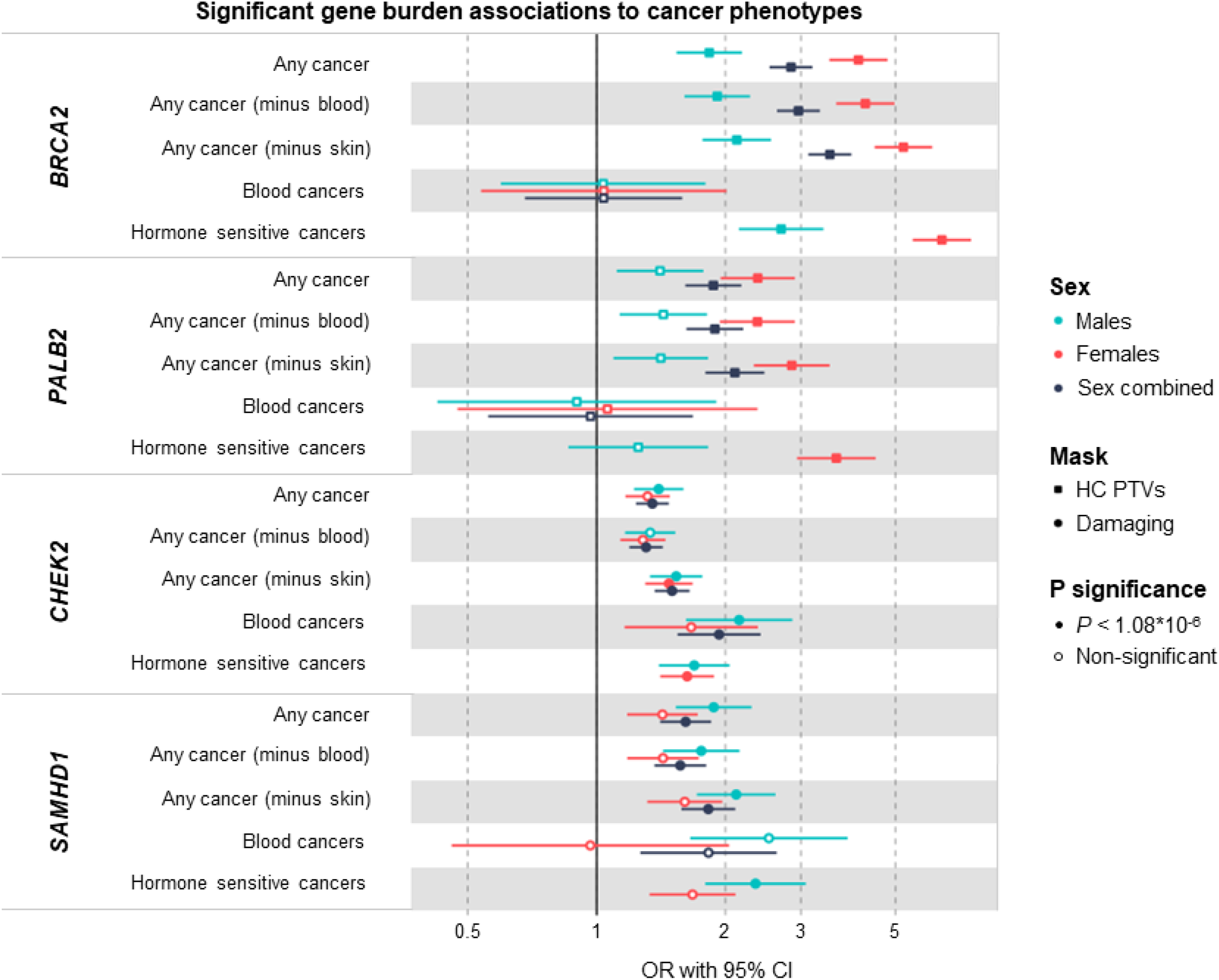
Forest plot for ANM WES genes with significant gene burden associations for cancer phenotypes. Exome-wide significant (P < 1.08*10^-6^) genes are displayed, showing sex-stratified and combined results. Hormone sensitive cancers were only tested in males and females separately (Methods). The presented masks were selected based on the most significant association per gene and cancer type. Points and bars indicate OR and 95% CI for specific genes and their variant categories in cancer. Filled symbols indicate a result passing a Bonferroni-corrected significance threshold of P < 1.08*10^-6^.

*SAMHD1* associations with cancer appear to be driven by increased risk for multiple site-specific cancers, notably prostate cancer in males, mesothelioma in both males and females, and suggestive evidence for higher breast cancer susceptibility in females (**Figure 4, Supplementary Table 11**). Although the numbers of mutation carriers diagnosed with each site-specific cancer was small, the majority of these findings persisted using logistic regression with penalised likelihood estimation, which is more robust to extreme case/control imbalance^34^ (**Supplementary Table 11**).

**Figure 4:**
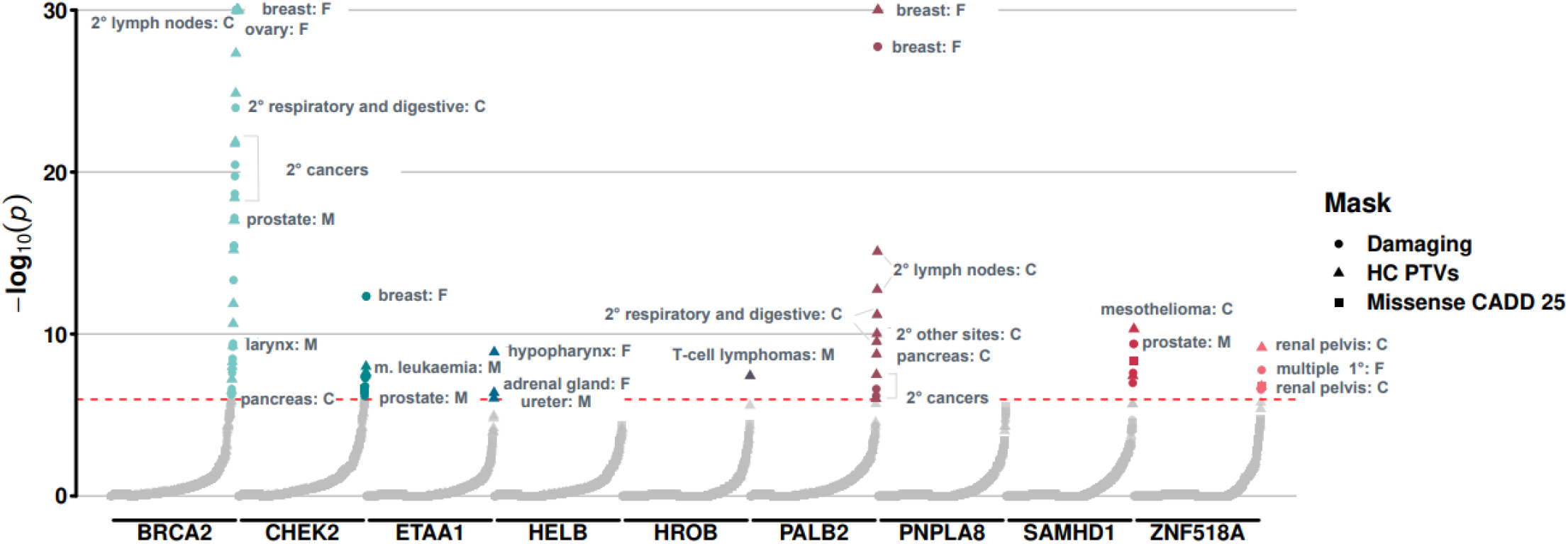
Genetic susceptibility to premature ovarian ageing and increased risk for diverse cancer types. Plot showing the association between loss of ANM genes identified in this study and risk of 90 site specific cancers among UK Biobank participants. Summary statistics for cancer associations were obtained using a logistic regression with penalised likelihood estimation that controls for case/control imbalance (Methods)^34^. Associations highlighted in text passed exome-wide significance (P < 1.08*10^-6^). The y-axis is capped at -log_10_(P) = 30 for visualisation purposes; un-capped summary statistics can be found in **Supplementary Table 11. F:** females, **M:** males, **C:** sex-combined. **1°:** primary cancer, **2°:** secondary cancer.

Cancer risk-increasing alleles in *SAMHD1* were associated with later ANM, which is similar to the pattern demonstrated previously for *CHEK2*. This finding is consistent with a mechanism of disrupted DNA damage sensing and apoptosis, resulting in slowed depletion of the ovarian reserve^1^. In addition, we provide robust evidence for a previously described rare variant association for *SAMHD1* with telomere length^35^, highlighting that rare damaging variants cause longer telomere length (*P*=1.4*10^-59^) (**Supplementary Table 10, Supplementary Figure 7**).

### Genetic susceptibility to ANM in mothers influences *de novo* mutation rate in offspring

Our previous common variant analyses demonstrated that many ANM associated variants implicate DNA damage repair (DDR) genes, an observation mirrored here in our rare variant associations. Therefore, we sought to test the hypothesis that inter-individual variation in these DDR processes would influence the mutation rate in germ cells and hence in the offspring. More specifically, we hypothesised that genetic susceptibility to earlier ovarian ageing would be associated with a higher *de novo* mutation (DNM) rate in the offspring. To test this, we analysed 8,089 whole-genome sequenced parent-offspring trios recruited in the rare disease programme of the 100,000 Genome Project (100kGP, **Supplementary Figure 8**). We calculated a polygenic score (PGS) for ANM in the parents based on our previously identified 290 common variants^1^ and tested this against the phased DNM rate in the offspring, adjusted for age. We found that maternal genetic susceptibility to earlier ANM was associated with an increased rate of maternally-derived DNMs in the offspring (rate ratio = 1.02 per SD of PGS, *P*=6.8*10^-4^, N=8,089 duos with European ancestry; **Supplementary Table 12**). We confirmed this finding in sensitivity analyses using the same data, in a two-sample Mendelian Randomization (MR) framework that can better model the dose-response relationship of these variants (**Supplementary Table 13**). These results were highly concordant, with all models showing a significant result and no heterogeneity (*P*min=6.3*10^-5^). In contrast, the paternal PGS was not associated with paternally-derived DNMs (*P*=0.51, N=8,029) nor was the maternal PGS associated with paternally-derived DNMs (*P*=0.55).

## Discussion

Our study extends the number of genes implicated in ovarian ageing through the identification of rare, protein-coding variants. Effect sizes ranged from 5.61 years earlier ANM for HC-PTV carriers in *ZNF518A*, to 1.35 years later ANM for women carrying damaging variants in *SAMHD1* compared to a maximum effect size of 1.06 years (median 0.12 years) reported for common variants (MAF>1%)^1^. Several of these effect estimates were comparable to those conferred by *FMR1* premutations, which are currently used as part of the only routinely applied clinical genetics test for premature ovarian insufficiency (POI)^36^. Deleterious variants in three genes (*CHEK2, HELB* and *SAMHD1*) were associated with an increase in ANM and therefore represent potential therapeutic targets for enhancing ovarian stimulation in women undergoing *in vitro* fertilisation (IVF) treatment through short-term apoptotic inhibition. Seven of the nine ANM genes identified have known roles in DNA damage repair, and three of these are linked to ANM for the first time (*PALB2, ETAA1* and *HROB*): *PALB2* is involved in *BRCA2* localization and stability and compound heterozygous mutations result in Fanconi anaemia and predispose to childhood malignancies^37^. *ETAA1* accumulates at DNA damage sites in response to replication stress^38–41^ and *HROB* is involved in homologous recombination by recruiting the *MCM8*-*MCM9* helicase to sites of DNA damage to promote DNA synthesis^42,43^. Homozygous loss-of-function of *HROB* is associated with POI^44^ and infertility in both sexes in mouse models^42^.

Novel biological mechanisms of ovarian ageing were revealed by finding associations with two non-DDR genes (*PNPLA8* and *ZNF518A*): *PNPLA8* is a calcium-independent phospholipase^45–47^ and a recessive cause of neurodegenerative mitochondrial disease and mitochondrial myopathy^48–52^; an association with reproductive phenotypes has not been described previously. *ZNF518A* belongs to the zinc finger protein family and is likely a transcriptional regulator for a large number of genes ^23^. We found that female carriers of rare protein truncating variants in *ZNF518A* have shorter reproductive lifespan due to delayed puberty timing and earlier menopause. Enrichment of GWAS signals at *ZNF518A* binding sites suggests that *ZNF518A* regulates the genes involved in reproductive longevity by repression of elements distal to transcription start sites.

While mutation in *SAMHD1* is a common somatic event in a variety of cancers^53–63^, it has not been described as a germline risk factor previously. Recessive inheritance of *SAMHD1* missense and PTV variants have been associated with Aicardi–Goutieres syndrome, a congenital autoimmune disease^64^. Our identified damaging variants in *SAMHD1* increased risk of ‘All cancer’ in males and females, as well as in sex-specific cancers, highlighting *SAMHD1* as a novel risk factor for prostate cancer in males and hormone-sensitive cancers in females. *SAMHD1* has a role in preventing the accumulation of excess deoxynucleotide triphosphates (dNTPs), particularly in non-dividing cells^65^. A regulated dNTP pool is important for the fidelity of DNA repair, thus highlighting additional roles of this gene in facilitation of DNA end resection during DNA replication and repair^65–70^. *SAMHD1* deficiency leads to resistance to apoptosis^71,72^, suggesting that delayed ANM might originate from slowed depletion of ovarian reserve due to disrupted apoptosis, analogous to the mechanism for *CHEK2* that has been reported previously.

Previous studies have demonstrated that parental age is strongly associated with the number of *de novo* mutations in offspring^73^, with the majority of these mutations arising from the high rate of spermatogonial stem cell divisions that underlie spermatogenesis throughout adult life of males^74^. Our current study provides the first direct evidence that maternal mutation rate is heritable, with women at higher genetic risk of earlier menopause transmitting an increased rate of *de novo* mutations to offspring. This could have direct implications for the health of future generations given the widely reported link between *de novo* mutations and increased risk of psychiatric disease and developmental disorders^75–78^. We speculate that if genetic susceptibility to earlier menopause influences *de novo* mutation rate, it is possible that non-genetic risk factors for earlier ANM, such as smoking and alcohol intake, would likely have the same effect^79^. Our observations makes conceptual sense given that menopause timing appears to be primarily driven by the genetic integrity of oocytes and their ability to sustain, detect, repair and respond to acquired DNA damage^1^. These observations also build on earlier work in mice and humans that *BRCA1*/*2* deficiency increases the rate of double strand breaks in oocytes and reduces ovarian reserve^80–82^.

An important limitation of our study, shared by many other similar large-scale exome sequencing studies, is that we were unable to replicate our findings in an independent cohort. Instead, we aimed to accumulate additional evidence where possible to support our observations and evaluate the biological plausibility of our findings. For example, the identified rare loss of function alleles in *ZNF518A* have the largest effect on ovarian ageing reported to date, which is supported by high expression in fetal germ cells, genome-wide significant common variants at the same locus, and the observation that *ZNF518A* binding sites genome-wide are significantly enriched for common variant ANM association(s). For all identified genes further experimental studies will ultimately be required to fully understand the biological mechanisms governing the observed effects on ovarian ageing.

## Methodology

### UK Biobank Data Processing and Quality Control

To conduct rare variant burden analyses described in this study, we obtained Whole Exome Sequencing data (WES) for 454,787 individuals from the UK Biobank study^83^. Participants were excluded based on excess heterozygosity, autosomal variant missingness on genotyping arrays ≥ 5%, or inclusion in the subset of phased samples as defined in Bycroft *et al*^84^. Analysis was restricted to participants with European genetic ancestry, leaving a total of 421,065 individuals. Variant quality control (QC) and annotation were performed using the UK Biobank Research Analysis Platform (RAP; https://ukbiobank.dnanexus.com/), a cloud-based central data repository for UK Biobank WES and phenotypic data. Besides the QC described by Backman *et al*.^83^, we performed additional steps using custom applets designed for the RAP. Firstly, we processed provided population-level Variant Call Format (VCF) files by splitting and left-correcting multi-allelic variants into separate alleles using ‘bcftools norm’^85^. Secondly, we performed genotype-level filtering applying ‘bcftools filter’ separately for Single Nucleotide Variants (SNVs) and Insertions/Deletions (InDels) using a missingness-based approach. Using this approach, we set to missing (i.e. ./.) all SNV genotypes with depth < 7 and genotype quality < 20 or InDel genotypes with a depth < 10 and genotype quality < 20. Next, we applied a binomial test to assess an expected alternate allele contribution of 50% for heterozygous SNVs; we set to missing all SNV genotypes with a binomial test p. value ≤ 1×10^-3^. Following genotype-level filtering we recalculated the proportion of individuals with a missing genotype for each variant and filtered all variants with a missingness value > 50%. The variant annotation was performed using the ENSEMBL Variant Effect Predictor (VEP) v104^86^ with the ‘--everything’ flag and plugins for CADD^87^ and LOFTEE^88^ enabled. For each variant we prioritised the highest impact individual consequence as defined by VEP and one ENSEMBL transcript as determined by whether or not the annotated transcript was protein-coding, MANE select v0.97, or the VEP Canonical transcript. Following annotation, variants were categorised based on their predicted impact on the annotated transcript. Protein Truncating Variants (PTVs) were defined as all variants annotated as stop gained, frameshift, splice acceptor, and splice donor. Missense variant consequences are identical to those defined by VEP. Only autosomal or chrX variants within ENSEMBL protein-coding transcripts and within transcripts included on the UKBB ES assay^83^ were retained for subsequent burden testing.

### Exome-wide association analyses in the UK Biobank

In order to perform rare variant burden tests, we used a custom implementation of BOLT-LMM v2.3.6^89^ for the RAP. Two primary inputs are required by BOLT-LMM: i) a set of genotypes with minor allele count > 100 derived from genotyping arrays to construct a null linear mixed effects model and ii) a larger set of variants collapsed on ENSEMBL transcript to perform association tests. For the former, we queried genotyping data available on the RAP and restricted to an identical set of individuals included for rare variant association tests. For the latter, and as BOLT-LMM expects imputed genotyping data as input rather than per-gene carrier status, we created dummy genotype files where each variant represents one gene and individuals with a qualifying variant within that gene are coded as heterozygous, regardless of the number of variants that individual has in that gene.

To test a range of variant annotation categories for MAF < 0.1%, we created dummy genotype files for high confidence PTVs as defined by LOFTEE, missense variants with CADD ≥ 25, and damaging variants that included both high confidence PTVs and missense variants with CADD ≥ 25. For each phenotype tested, BOLT-LMM was then run with default parameters other than the inclusion of the ‘lmmInfOnly’ flag. To derive association statistics for individual markers, we also provided all 26,657,229 individual markers regardless of filtering status as input to BOLT-LMM. All tested phenotypes were run as continuous traits corrected by age, age^2^, sex, the first ten genetic principal components as calculated in Bycroft *et al*^84^ and study participant ES batch as a categorical covariate (either 50k, 200k, or 450k).

For discovery analysis in the primary trait of interest, age at natural menopause, we analysed 17,475 protein-coding genes with the minimum of 10 rare allele carriers in at least one of the masks tested using BOLT-LMM (**Supplementary Table 1**). The significant gene-level associations for ANM were identified applying Bonferroni correction for the number of masks with MAC≥10 (N=46,251 masks) in 17,475 protein-coding genes (*P*: 0.05/46,251 = 1.08*10^-6^) (**Supplementary Table 2**).The age at natural menopause results obtained via BOLT-LMM are available in **Supplementary Table 1**. Furthermore, in order to compare and explain potential differences between our WES results and the previously published one^9^, we ran the above described approach using MAF < 1%, a cutoff applied by Ward *et al*. (**Supplementary Table 3, Supplementary Note**).

To generate accurate odds ratio and standard error estimates for binary traits, we also implemented a generalised linear model using the statsmodels package^90^ for python in a three step process. First, a null model was run with the phenotype as a continuous trait, corrected for control covariates as described above. Second, we regressed carrier status for individual genes on the residuals of the null model to obtain a preliminary *P* value. Thirdly, all genes were again tested using a full model to obtain odds ratios and standard errors with the family set to ‘binomial’. Generalised linear models utilised identical input to BOLT-LMM converted to a sparse matrix.

### Phenotype derivation

Age at natural menopause was derived for individuals within the UK Biobank, who were deemed to have undergone natural menopause, i.e. not affected by surgical or pharmaceutical interventions, as follows:

Firstly, European female participants (n=245,820) who indicated during any of the attended visits having had a hysterectomy were collated (fields 3591 and 2724) and their reported hysterectomy ages were extracted (field 2824) and the median age was kept (n=47,218 and 46,260 with reported ages). The same procedure was followed for participants indicating having undergone a bilateral oophorectomy (surgery field 2834 and age field 3882, n=20,495 and 20,001 with reported ages).

For individuals having indicated the use of hormone replacement therapy (HRT; field 2814), HRT start and end ages were collated (fields 3536 and 3546, accordingly) across the different attended visits (n=98,104). In cases where the reported chronological HRT age at later attended visits was greater than that at previous visits, the later instances were prioritised, i.e. as they would potentially indicate an updated use of HRT. In cases where different HRT ages were reported, but not in chronologically increasing order, the median age was kept.

Menopausal status was determined using data across instances (field 2724) and prioritising the latest reported data, to account for changes in menopause status. For participants indicating having undergone menopause, their reported ages at menopause were collated (field 3581) using the same procedure as for HRT ages (n=158,264).

Exclusions were then applied to this age at menopause, as follows:

- Participants reporting undergoing a hysterectomy and/or oophorectomy, but not the age at which this happened (n=958 and 494, accordingly)
- Participants reporting multiple hysterectomy and/or oophorectomy ages, which were more than 10 years apart (n=38 and 23, accordingly)
- Participants reporting multiple HRT start and/or end ages, which were not in chronologically ascending order and were more than 10 years apart (n=124 and 137, accordingly)
- Participants reporting multiple ages at menopause, which were not in chronologically ascending order and were more than 10 years apart (n=73) and participants who reported both having and not having been through menopause and no other interventions (n=98)
- Participants having undergone a hysterectomy/oophorectomy before or during the year they report undergoing menopause
- Participants starting HRT prior to undergoing menopause and participants reporting HRT use, with no accompanying dates

The resulting trait was representative of an age at natural menopause (ANM, n=115,051) and was used in downstream analyses. Two additional ANM traits were also calculated, windsorized one by coding everyone reporting an ANM younger than 34, as 34 used in the discovery analysis as the primary phenotype (n=115,051 total, reduced to 106,973 after covariate-resulting exclusions), and one by only including participants reporting ANM between 40 and 60, inclusive (n=104,506), treated as a sensitivity analysis.

All manipulations were conducted in R (v4.1.2) on the UKB Research Analysis Platform (RAP; https://ukbiobank.dnanexus.com/).

### Phenome-wide association analysis

In order to test the association of ANM identified genes in other phenotypes, we processed additional reproductive ageing-related phenotypes, including age at menarche, cancer, telomere length (TL) and sex hormones (SH). All tested phenotypes were run as either continuous (age at menarche, TL and SH) or binary traits (cancer) corrected by age, age^2^, sex, the first ten genetic principal components as calculated in Bycroft *et al*^84^, and study participant ES batch as a categorical covariate (either 50k, 200k, or 450k). Phenotype definitions and processing used in this study are described in **Supplementary Tables 8 and 9**. Only the first instance (initial visit) was used for generating all phenotype definitions unless specifically noted in **Supplementary Table 8**. In case of cancer-specific analysis data from cancer registries, death records, hospital admissions and self-reported were harmonised to ICD10 coding. If a participant had a code for any of the cancers recorded in ICD10 (C00-C97) then they were counted as a case for this phenotype. Minimal filtering was performed on the data, with only those cases where a diagnosis of sex-specific cancer was given in contrast to the sex data contained in UK Biobank record 31, was a diagnosis not used. For more information on cancer specific analysis refer to **Supplementary Tables 9 and 11**.

### Cancer PheWAS Associations

To test for an association between genes we identified as associated with menopause timing (**Supplementary Table 2, Figure 1**) and 90 individual cancers as included in cancer registries, death records, hospital admissions and self-reported data provided by UK Biobank (e.g. breast, prostate, etc.) we utilised a logistic model with identical covariates as used during gene burden testing (N = 2430 tests) (**Supplementary Tables 9 and 11**). As standard logistic regression can lead to inflated *P* value estimates in cases of severe case/control imbalance^91^, we also performed a logistic regression with penalised likelihood estimation as described by Firth^34^ (**Supplementary Table 12**). Models were run as discussed in Kosmidis *et al*.^92^ using the ‘brglm2’ package implemented in R. brglm2 was run via the ‘glm’ function with default parameters other than “family” set to “binomial”, “method” set to “brglmFit”, and “type” set to “AS_mean”.

### WES sensitivity analysis using REGENIE

To replicate the primary findings and account for potential bias that could be introduced by exclusively using one discovery approach, a second analyst independently derived the age at menopause phenotype using a previously published method^93^ and conducted additional burden association analysis using the REGENIE regression algorithm (REGENIEv2.2.4; https://github.com/rgcgithub/regenie). REGENIE implements a generalised mixed-model region-based association test that can account for population stratification and sample relatedness in large-scale analyses. REGENIE runs in 2 steps^94^, which we implemented on the UKBiobank RAP: In the first step, genetic variants are aggregated into gene specific units for each class of variant called masks. We selected variants in CCDS transcripts deemed to be high confidence by LOFTEE^88^ with MAF<0.1% and annotated using VEP^86^. We created three masks, independently of primary analysis group: (1) loss-of-function (LOF) variants (stop-gain, frameshift, or abolishing a canonical splice site (-2 or +2 bp from exon, excluding the ones in the last exon)) or missense variants with CADD score >30, (2) LOF or missense variants with CADD score >25, (3) all missense variants. In the second step, the three masks were tested for association with ANM. We applied an inverse normal rank transformation to ANM and included recruitment centre, sequence batch and 40 principal components as covariates. For each gene, we present results for the transcript with the smallest burden *P* value. The results for the sensitivity analysis performed via REGENIE are available in **Supplementary Table 1**.

### Common variant GWAS lookups

Genes within 500kb upstream and downstream of the 290 lead SNPs from the latest GWAS of ANM^1^ were extracted from the exome-wide analysis. There were a total of 2149 genes within the GWAS regions. Burden tests in these genes with a Bonferroni corrected *P* value of <2.3*10^-5^ (0.05/2149) were highlighted. The results are available in **Supplementary Table 4**.

### Analysis of GWAS and WES genes expression profiles in human female germ cells at various stages of development

We studied the mRNA abundance of WES genes during various stages of human female germ cell development using single-cell RNA sequencing data (**Supplementary Tables 6 and 7**). We used the processed single cell RNA resequencing datasets from two published studies. This included single-cell RNA sequencing data from foetal primordial germ cells of human female embryos (Accession code: GSE86146^95^), and from oocyte and granulosa cell fractions during various stages of follicle development (Accession code: GSE107746^96^). A pseudo score of 1 was added to all values before log transformation of the dataset. The samples from fetal germ cells (FGCs) were categorised into sub-clusters as defined in the original study. The study by Li *et al*^95^ had identified 17 clusters by performing a t-distributed stochastic neighbour embedding (t-SNE) analysis and using expression profiles of known marker genes for various stages of fetal germ cell development. In our analysis we have included four clusters of female FGCs (Mitotic, Retinoic Acid (RA) responsive, Meiotic, Oogenesis) and four clusters containing somatic cells in the fetal gonads (Endothelial, Early_Granulosa, Mural_Granulosa, Late_Granulosa). Software packages for R - tidyverse (https://www.tidyverse.org/), pheatmap, (https://CRAN.R-project.org/package=pheatmap), reshape2 (https://github.com/hadley/reshape), were used in processing and visualising the data.

### Functional enrichment tests for ZNF518A transcription factor binding sites using fGWAS and SLDP

fGWAS (*v*.*0*.*3*.*6)*, a hierarchical model for joint analysis of GWAS and genomic annotations, was implemented to test the functional enrichment of ANM GWAS hits in ZNF518A transcription factor binding sites^26^. The fGWAS input file contained the ANM GWAS summary stats derived from the Reprogen study^1^ annotated for *ZNF518A* binding sites. The *ZNF518A* annotation file was derived from the ENCODE ChIP-seq data from human HEK293 cell line^97^ the optimal independent discovery rate peak calling against hg19 [ENCFF415VBF] was used. The ANM GWAS hits were annotated for the presence/absence of the *ZNF518A* transcription factor binding sites in a binary way (0, 1), with ‘1’ if the SNP falls within the transcription factor binding site and ‘0’ otherwise. The fGWAS tool available from https://github.com/joepickrell/fgwas and was run in annotation mode “-w” for the describe *ZNF518A* annotation. Detailed description of fGWAS methodology is available in Pickrell *et al*, 2014^26^. In short, the genome is split into independent blocks, which are allowed to contain either a single polymorphism that causally influences the trait or none. fGWAS then models the prior probability that any given block contains an association and the conditional prior probability that any given SNP in the block is the causal one, with probabilities allowed to vary according to functional annotations. The priors are then estimated using an empirical Bayes approach. The fGWAS output contained the maximum likelihood parameter estimates for each parameter in the model, in this case *ZNF518A*, with the lower and upper bound of the 95% confidence interval (CI) on the parameter. The P value was calculated from lower and upper CI in 3 following steps: (1) Standard error (SE) calculation: ***SE* = (*Upper CI* - *Lower CI*)/(2*1.96)**; (2) Test statistics calculation: ***Z=Estimate / SE***; and (3) P value calculation: ***P = exp(-0*.*717*Z - 0*.*416*Z***^***2***^).

Signed LD profile (SLDP) regression was applied to explore the directional effect of a signed functional annotation, *ZNF518A*, on a heritable trait like ANM using GWAS summary statistics. More specifically, we tested whether alleles that are predicted to increase the binding of the transcription factor *ZNF518A* have a genome-wide tendency to increase or decrease timing of menopause in women. The SLDP tool was installed from https://github.com/yakirr/sldp, with the comprehensive methodological steps described in Reshef *et al*, 2018^27^. For the analysis to be conducted, SLDP required GWAS summary statistics for ANM, signed LD profiles for *ZNF518A* binding, signed background model and reference panel in a SLDP compatible format. For the reference we used a 1000 Genomes Phase 3 European reference panel in *plink* format, which contained approximately 10M SNPs and 500 people and was available for download at the <u>‘refpanel’</u> page. The ANM GWAS summary statistics, available from our latest Reprogen study^1^, was pre-processed using the ‘*preprocesspheno*’ tool from the SLDP package. To conduct this step, we also obtained the list of regression SNPs along with the LD scores for the reference panel from the <u>‘refpanel’</u> page. The pre-processing step included filtering down to SNPs that are also present in the reference panel, harmonising alleles to the reference, and multiplying the summary statistics by the SLDP regression weights. In addition, we applied the ‘*preprocessrefpanel*’ tool to compute a truncated singular value decomposition (SVD) for each LD block in the reference panel. These SVDs were later used to weight the SLDP regression. The *ZNF518A* annotation file was obtained from the ENCODE CHIP-seq analysis, as described above, and preprocessed using the ‘*preprocessannot*’ tool that turns signed functional annotations into signed LD profiles. Prior to running SLDP, we also obtained the signed background LD profiles that enabled us to control for systematic signed effects of minor alleles, which could arise from either population stratification or negative selection. SLDP was then run on our data using ‘*sldp*’ function. To explore the relevance of *ZNF518A* for menopause timing in comparison to other transcription regulators, we tested whether genome-wide sequence changes introduced by SNP alleles identified in ANM GWAS increase or decrease binding of additional 382 transcription factors (TFs). The preprocessed annotation files for 382 TFs derived from ENCODE CHIP-seq experiments, were available for download at the <u>annotation data page</u>. The results are available in **Supplementary Table 5**.

### Functional analysis of ZNF518a binding sites

*ZNF518A* peaks were derived from unique genomic regions in ENCODE accession ENCFF415VBF described above. Quantification of ChIP-seq signal by aligning paired-end replicates (ENCFF174HBR, ENCFF574GQY, ENCFF808AJP, ENCFF453FDD) to the hg19 genome with Bowtie2 v2.3.5.1^98^ with options “-I 0 -X 1000 –no-discordant –no-mixed”, reads were filtered for those with MAPQ > 30 with samtools v1.10. Assessment of H3K27ac^29^ and chromatin accessibility by ATAC-seq^30^ in day 4 human primordial germ cell like cells (hPGCLCs) at *ZNF518A* peaks was performed. For H3K27ac single end reads from accessions GSM4257216, GSM4257217, GSM4257218 were obtained and aligned with Bowtie2 v2.3.5.1 with default settings and MAPQ > 30 reads retained as above. For ATAC-seq paired-end reads were obtained from accessions GSM3406938, GSM3406939 and mapped and filtered as *ZNF518A* reads above.

Quantification of ChIP-seq and ATAC-seq signals for peak heights, heatmaps was performed with https://github.com/owensnick/GenomeFragments.jl. Peak to TSS distances were calculated against Gencode v36 release liftover to hg19 using GenomicFeatures.jl and https://github.com/owensnick/ProximityEnrichment.jl. We consider four categories of peaks: TSS intersecting, TSS proximal (TSS < 2000kb, outside gene body), Gene body intersecting, Intergenic and Distal (TSS > 5kb).

To perform *de novo* motif discovery we used Homer v4.11.1^99^ using findMotifsGenome.pl with options “hg19 -size 200”. We ran this on all ZNF518A peaks, distal peaks and those intersecting TSS, we recovered a motif matching JASPAR^28^ unvalidated motif UN0199.1 in all peak sets apart from those intersecting TSS. We then used https://github.com/exeter-tfs/MotifScanner.jl to quantify the occurrence of all instances of motif UN0199.1 in ZNF518A peaks.

We downloaded the 18-state ChromHMM^100^ models for all 833 biosamples in Epimap^31^ from http://compbio.mit.edu/epimap/. We calculated the intersection between each state in each biosample and either all ZNF518A peaks or distal ZNF518A peaks using GenomicFeatures.jl. We calculated odds ratios from contingency tables using the approximation of bedtools^101^ and Giggle^102^, by estimating total genomic intervals as hg19 genome size divided by the sum of the mean ZNF518A peak size and the chromatin state interval size.

### *De novo* mutation rate analyses

We calculated polygenic scores (PGSs) in participants from the rare disease programme of the 100,000 Genome Project (100kGP) v14. There are 77,901 individuals in the Aggregated Variant Calls (aggV2) after excluding participants whose genetically inferred sex is not consistent with their phenotypic sex. We restricted the PGS analysis to individuals of European ancestry, which was predicted by the Genomics England Bioinformatics team using a random forest model based on genetic principal components (PCs) generated by projecting aggV2 data onto the 1000 Genomes phase 3 PC loadings. We removed one sample in each pair of related probands with kinship coefficient > 1/(2^4.5), i.e. up to and including third degree relationships. Probands with the highest number of relatives were removed first. Similarly, we retained unrelated mothers and fathers of these unrelated probands. It left us with 8,089 mother-offspring duos and 8,029 father-offspring duos.

We used the lead variants (or proxies, as described below) for genome-wide significant loci previously reported for ANM^1^ to calculate PGS in the parents. In 100kGP, we removed variants with minor allele frequency (MAF) <0.5% or missing rate >5% from the aggV2 variants prepared by the Genomics England bioinformatics team. For lead variants that did not exist in 100kGP, we used the most significant proxy variants with linkage disequilibrium (LD) r^2^ >0.5 if available in 100kGP. This resulted in a PGS constructed from 287 of the 290 previously reported loci. We regressed out 20 genetic PCs that were calculated within the European subset from the PGS and scaled the residuals to have mean = 0 and standard deviation = 1. Higher PGS indicates later age at menopause.

De novo mutations (DNMs) were called in 10,478 parent offspring trios by the Genomics England Bioinformatics team. The detailed analysis pipeline is documented at: https://research-help.genomicsengland.co.uk/display/GERE/De+novo+variant+research+dataset. Extensive quality control (QC) and filtering were applied by Kaplanis *et al*. as described previously^103^. De novo single nucleotide variants (dnSNVs) were phased using a read-based approach based on heterozygous variants near the DNM that were able to be phased to a parent. About one third of the dnSNVs were phased, of which three quarters were paternally phased (**Supplementary Figure 8, Supplementary Table 12**).

In association models, we accounted for parental age, the primary determinant of the number of DNMs, and various data quality metrics as described in^103^:

- Mean coverage for the child, mother and father (child_mean_RD, mother_mean_RD, father_mean_RD)
- Proportion of aligned reads for the child, mother and father (child_prop_aligned, mother_prop_aligned, father_prop_aligned)
- Number of SNVs called for child, mother and father (child_SNVs, mother_SNVs, father_SNVs)
- Median variant allele fraction of DNMs called in child (median_VAF)
- Median ‘Bayes Factor’ as outputted by Platypus for DNMs called in the child. This is a metric of DNM quality (median_BF).

We first tested the association between parental PGSs and total dnSNV count in the offspring in a Poisson regression:

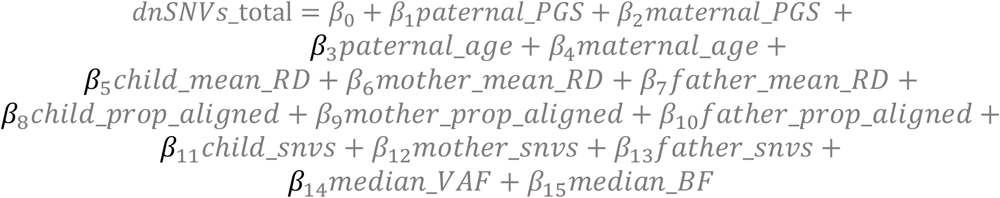

We also fitted Poisson regression models to test the association between the PGS of one of the parents and the dnSNVs in the offspring that were phased to the relevant parent.

The paternal model included paternal PGS, age, and data quality metrics that are related to the proband and the father:

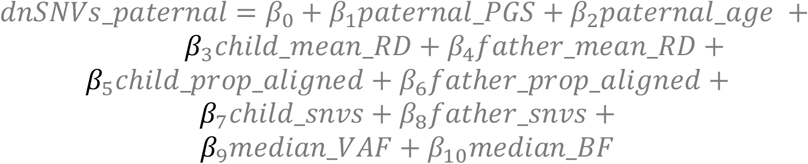

Similarly, the maternal model was as follows:

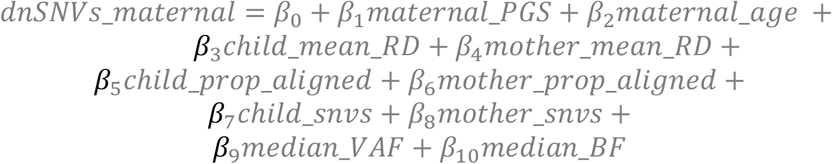

Finally, as a sanity check, we assessed the association between the maternal PGS and paternally phased dnSNVs, and vice versa:

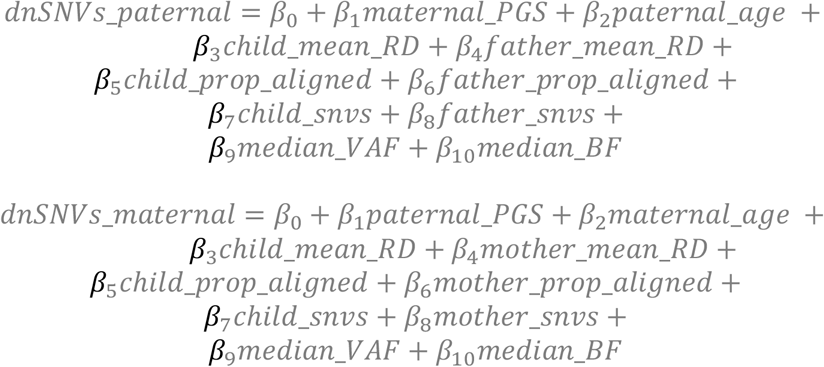

## Mendelian Randomization

### Instrumental variable selection

MR analysis was applied to examine the likelihood of a causal effect of polygenic score (PGS) of age at natural menopause on the risk of de novo mutation rates in the offspring (**Supplementary Table 13**). In this approach, genetic variants that are significantly associated with an exposure of interest are used as instrumental variables (IVs) to test the causality of that exposure on the outcome of interest^104–106^. For a genetic variant to be a reliable instrument, the following assumptions should be met: (1) the genetic instrument is associated with the exposure of interest, (2) the genetic instrument should not be associated with any other competing risk factor that is a confounder, and (3) the genetic instrument should not be associated with the outcome, except via the causal pathway that includes the exposure of interest^104,107^. Genotypes at all variants were aligned to designate the ANM PGS-increasing alleles as the effect alleles as described above and this was used as a genetic instrument of interest. The effect sizes of genetic instruments (genotypes in the mother) on maternally phased de novo SNVs in the offspring estimated in 8,089 duos were obtained from Genomics England.

### MR Frameworks

The MR analysis was conducted using the inverse-variance weighted (IVW) model as the primary model due to the highest statistical power^108^. However, as it does not correct for heterogeneity in outcome risk estimates between individual variants^109^, we applied a number of sensitivity MR methods that better account for heterogeneity^110^. These include MR Egger to identify and correct for unbalanced heterogeneity (‘horizontal pleiotropy’), indicated by a significant Egger intercept (*P*<0.05)^111^, and weighted median (WM) and penalised weighted median (PWM) models to correct for balanced heterogeneity^112^. In addition, we introduced the MR Radial method to exclude variants from each model in cases where they are recognized as outliers^113^. The results were considered as significant based on the *P* value significance consistency across different primary and sensitivity models applied. The results are available in **Supplementary Table 13**. Finally, in order to calculate the effect of ANM on offspring de novo mutation rate when comparing women with ANM at two extremes of the ANM distribution curve, we multiplied the effect obtained by MR IVW, i.e. a de novo count beta per 1 year change in ANM, by 20, an arbitrary number that compares women with ANM 20 years apart.

## Data Availability

All data produced using the UK Biobank resource will be returned to the UK Biobank returns catalogue upon publication.

## Acknowledgements

This work was funded by the Medical Research Council (Unit programs: MC_UU_12015/2, MC_UU_00006/2, MC_UU_12015/1, and MC_UU_00006/1). For the purpose of open access, the author has applied a Creative Commons Attribution (CC BY) licence to any Author Accepted Manuscript version arising. This research was conducted using the UK Biobank Resource under application 9905 (University of Cambridge) and 9072 and 871 (University of Exeter).

Ajuna Azad and Eva Hoffmann were supported by the ERC (724718-ReCAP), Novo Nordisk Foundation (NNF15COC0016662), the Independent Research Foundation Denmark (0134-00299B), and a grant from the Danish National Research Foundation Centre (6110-00344B).

Saleh Shekari was supported by the QUEX Institute (University of Exeter, UK and the University of Queensland, Australia). Anna Murray, Caroline Wright and Michael Weedon are supported by the Medical Research Council (MR/T00200X/1).The authors acknowledge the use of the University of Exeter High-Performance Computing facility in carrying out this work, funded by a MRC Clinical Research Infrastructure award (MRC Grant: MR/M008924/1).

This research was made possible through access to the data and findings generated by the 100,000 Genomes Project. The 100,000 Genomes Project is managed by Genomics England Limited (a wholly owned company of the Department of Health and Social Care). The 100,000 Genomes Project is funded by the National Institute for Health Research and NHS England. The Wellcome Trust, Cancer Research UK and the Medical Research Council have also funded research infrastructure. The 100,000 Genomes Project uses data provided by patients and collected by the National Health Service as part of their care and support.”

## Extended authorship list

Ambrose, J. C.^8^; Arumugam, P.^8^; Bevers, R.^8^; Bleda, M.^8^; Boardman-Pretty, F.^8,9^; Boustred, C. R.^8^; Brittain, H.^8^; Brown, M.A.; Caulfield, M. J.^8,9^; Chan, G. C.^8^; Giess A.^8^; Griffin, J. N.; Hamblin, A.^8^; Henderson, S.^8,9^; Hubbard, T. J. P.^8^; Jackson, R.^8^; Jones, L. J.^8,9^; Kasperaviciute, D.^8,9^; Kayikci, M.^8^; Kousathanas, A.7?; Lahnstein, L.^8^; Lakey, A.; Leigh, S. E. A.^8^; Leong, I. U. S.^8^; Lopez, F. J.^8^; Maleady-Crowe, F.^8^; McEntagart, M.^8^; Minneci F.^8^; Mitchell, J.^8^; Moutsianas, L.^8,9^; Mueller, M.^8,9^; Murugaesu, N.^8^; Need, A. C.^8,9^; O‘Donovan P.^8^; Odhams, C. A.^8^; Patch, C.^8,9^; Perez-Gil, D.^8^; Pereira, M. B.^8^; Pullinger, J.^8^; Rahim, T.^8^; Rendon, A.^8^; Rogers, T.^8^; Savage, K.^8^; Sawant, K.^8^; Scott, R. H.^8^; Siddiq, A.^8^; Sieghart, A.^8^; Smith, S. C.^8^; Sosinsky, A.^8,9^; Stuckey, A.^8^; Tanguy M.^8^; Taylor Tavares, A. L.^8^; Thomas, E. R. A.^8,9^; Thompson, S. R.^8^; Tucci, A.^8,9^; Welland, M. J.^8^; Williams, E.^8^; Witkowska, K.^8,9^; Wood, S. M.^8,9^; Zarowiecki, M.^8^.

