## Supplementary Figures for "Genetic susceptibility to earlier ovarian ageing increases *de novo* mutation rate in offspring"

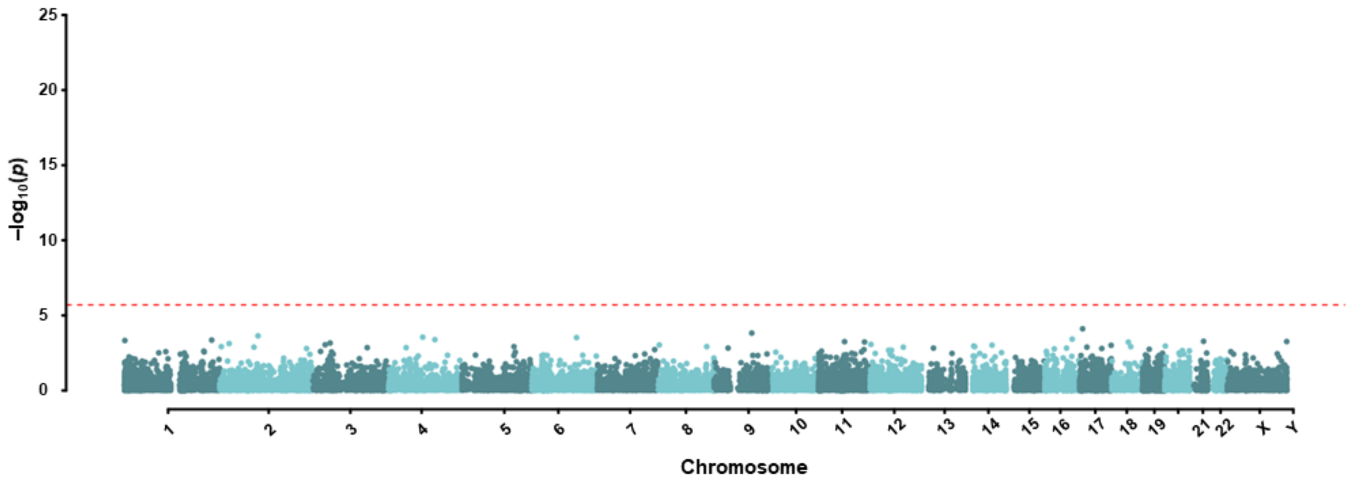


***Supplementary Figure 1: Exome-wide association results for synonymous variants.*** *Plotted are per-gene burden results for synonymous variants. The red line indicates the exome-wide significant P value after Bonferroni correction of $1.08*10^{-6}$.*

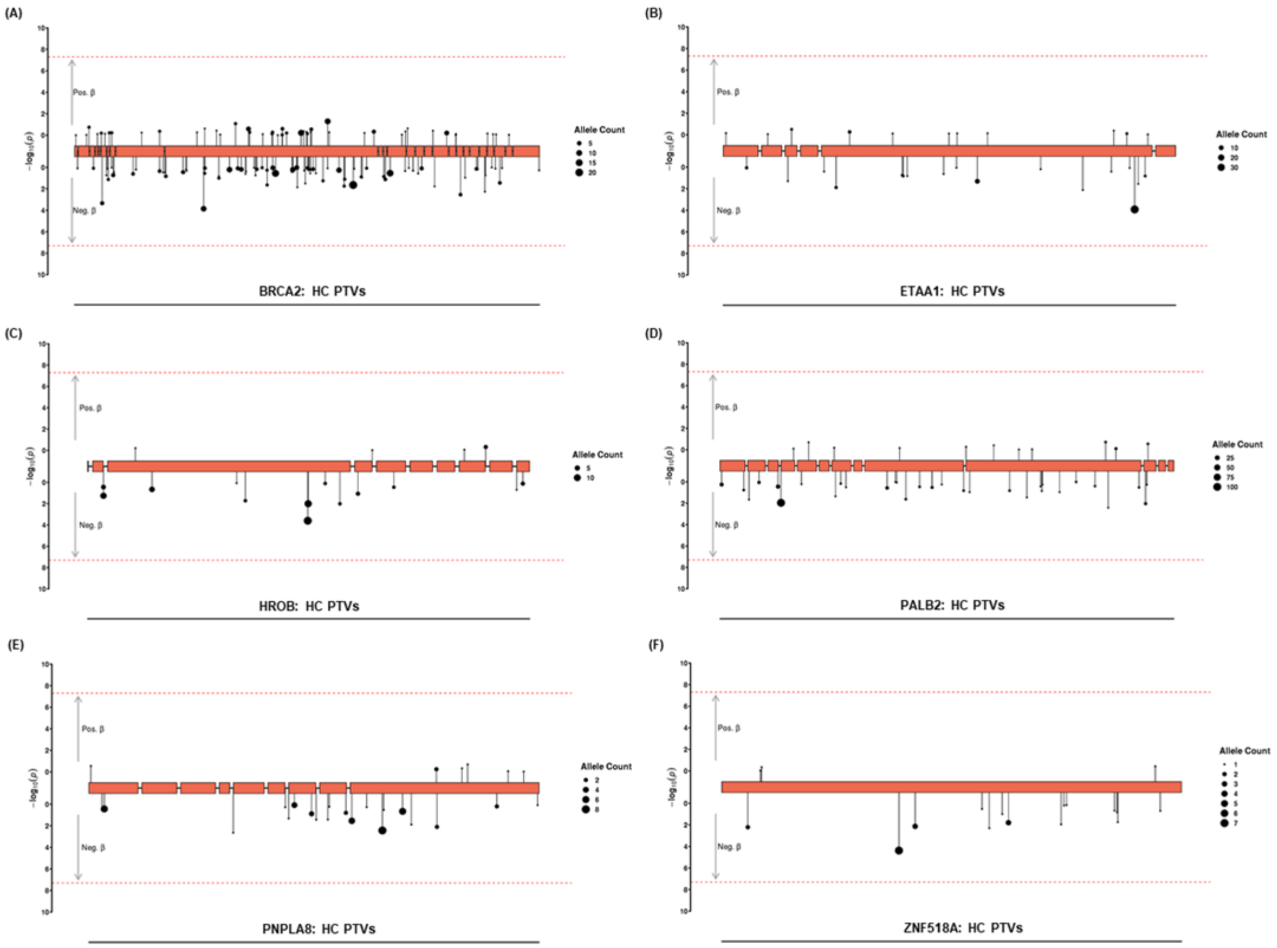


***Supplementary Figure 2: Variant level associations for ANM decreasing WES genes.*** *Lolliplot plots show the variants clustered for the best performing functional mask in a gene that went into the gene burden test for ANM using BOLT-LMM. These include:* ***(A)*** *BRCA2 HC PTV mask;* ***(B)*** *ETAA1, HC PTV mask;* ***(C)*** *HROB, HC PTV mask;* ***(D)*** *PALB2, HC PTV mask;* ***(E)*** *PNPLA8, HC PTV mask; and* ***(F)*** *ZNF518A, HC PTV mask. The arrows pointing upwards represent the variants positively associated with ANM, while the downwards ones show the negatively associated variants. The size of the point indicates the allele count in carriers.*

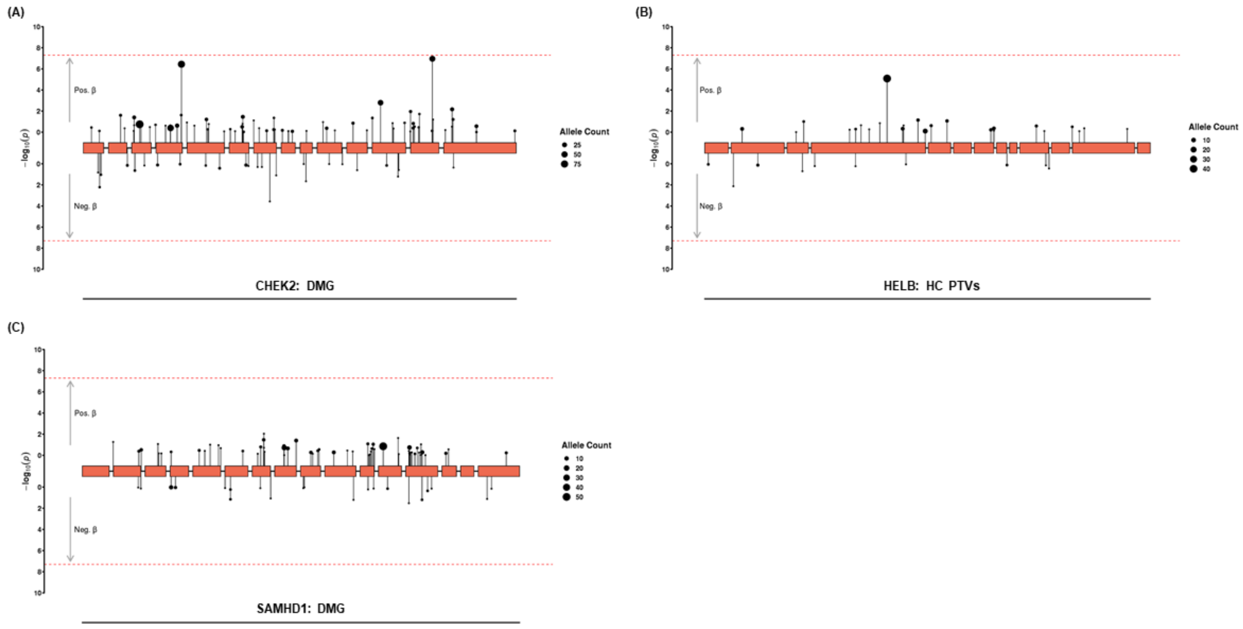


***Supplementary Figure 3: Variant level associations for ANM increasing WES genes.*** *Lolliplot plots show the variants clustered for the best performing functional mask in a gene that went into the gene burden test for ANM using BOLT-LMM. These include:* ***(A)*** *CHECK2, damaging mask;* ***(B)*** *HELB, HC PTV mask; and* ***(C)*** *SAMHD1, damaging mask. The arrows pointing upwards represent the variants positively associated with ANM, while the downwards ones show the negatively associated variants. The size of the point indicates the allele count in carriers.*

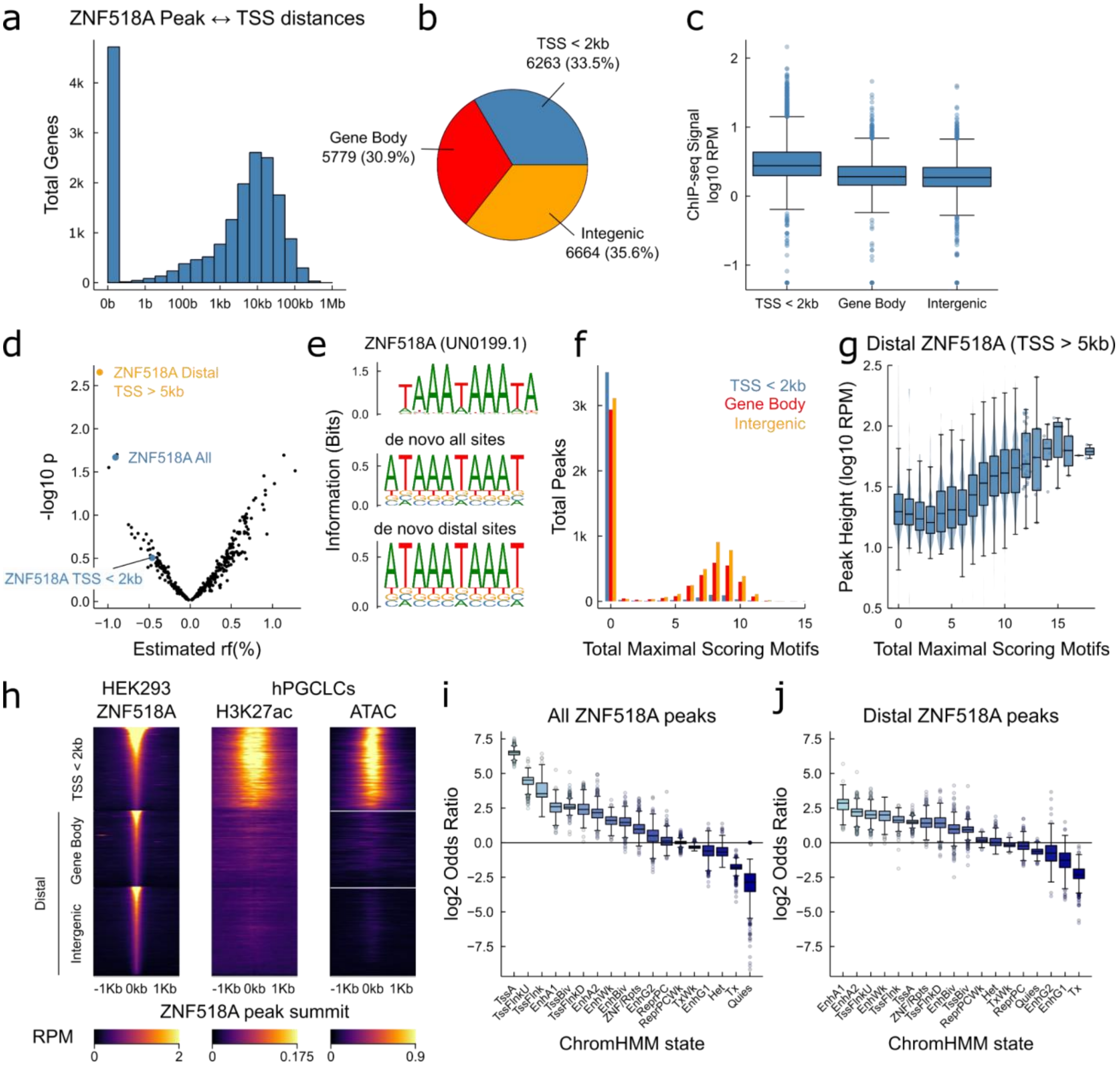


***Supplementary Figure 4: Functional analysis of ZNF518A bound loci. (a)*** *Histogram of log10-scale distances between ZNF518A and nearest gene transcription start site (TSS).* ***(b)*** *Proportion of ZNF18A peaks falling proximal to TSS (TSS < 2kb), within gene bodies and in intergenic regions.* ***(c)*** *Boxplots showing total normalised reads per million (RPM) for every peak for categories TSS < 2kb, gene body and intergenic - ZNF518A peaks have greater signal at proximal to TSS.* ***(d)*** *SLDP association between ANM GWAS variants and ZNF518A peaks, stratified by all peaks, proximal (< 2kb) from a TSS, and distal (> 5kb) from a TSS. The association between ANM variants and ZNF518A peaks appears due to distal ZNF518A peaks (either gene body or intergenic, > 5kb TSS) and not proximal TSS binding. Numerical results are reported in Supplementary Table 5.* ***(e)*** *De novo motif discovery recovers unvalidated JASPAR motif for ZNF518A UN0199.1. Homer enrichment statistics: all sites P= $10^{-6451}$ motif in 31.2% of targets (1.15% background); distal sites P= $10^{-4590}$ motif in 47.3% of targets (1.81 % background).* ***(f)*** *Proportion of maximal scoring instances of UN0199.1 (sequences that exactly match motif consensus) by ZNF518A peak category. Many distal peaks contain multiple perfect instances of the motif.* ***(g)*** *Boxplots, violin plots and dot plots depicting the relationship between ZNF518A ChIP-seq peak height and number of maximal scoring motifs present in peak. A strong relationship between peak height and number of motif instances can be observed.* ***(h)*** *Heatmaps depicting ZNF518A ChIP-seq, H3K27ac ChIP-seq in hPGCLCs, and*

*chromatin accessibility by ATAC-seq in hPGCLCs. Signal shown over all ZNF518A peaks in RPM +/- 1kb of ZNF518A peak summit. ZNF518A bound promoters (TSS < 2kb) are accessible and are marked with H3K27ac, distal regions either in gene bodies or intergenic regions show no H3K27ac or chromatin accessibility, suggestive that ZNF518A represses these regulatory regions.* ***(i,j)*** *Association shown in odds ratios of ChromHMM states over 833 tissues/cell types from Epimap, boxplots with outliers shown, each boxplot summarises the distribution of associations over all tissues/cell types for a given chromatin state.* ***(i)*** *All ZNF518A peaks;* ***(j)*** *ZNF518A peaks distal from TSS.*

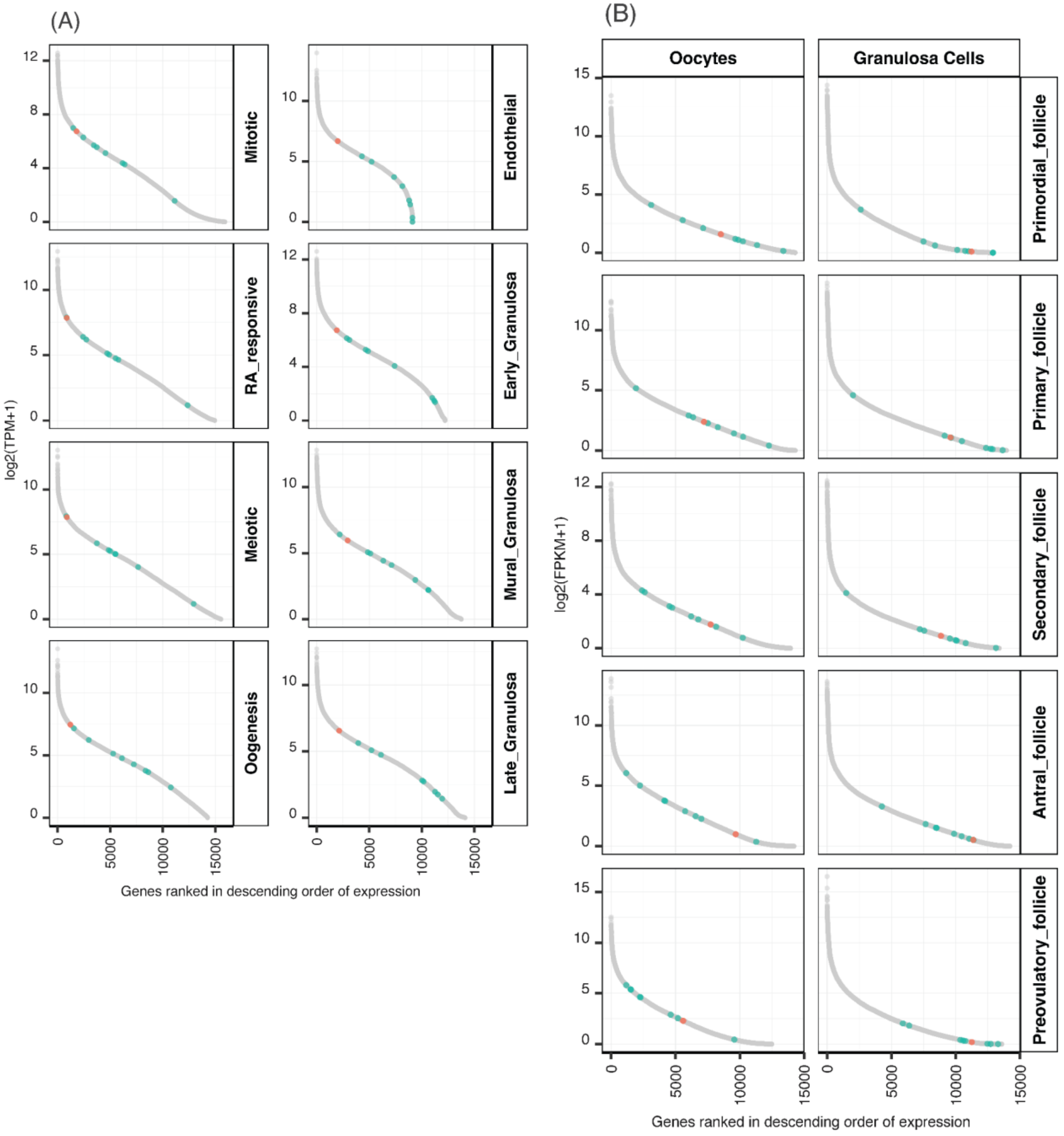


***Supplementary Figure 5: Expression levels of genes across various stages of female germ cell development.*** *In the X-axis, genes are ranked according to their average expression at each stage (Y-axis)* ***(A)*** *in human foetal primordial germ cells and* ***(B)*** *in granulosa cells in adult follicles. Genes identified as novel ANM genes in WES analysis are coloured in green and all other genes in the genome are in grey. ZNF518A is depicted in orange for the ease of comparison with other genes.*

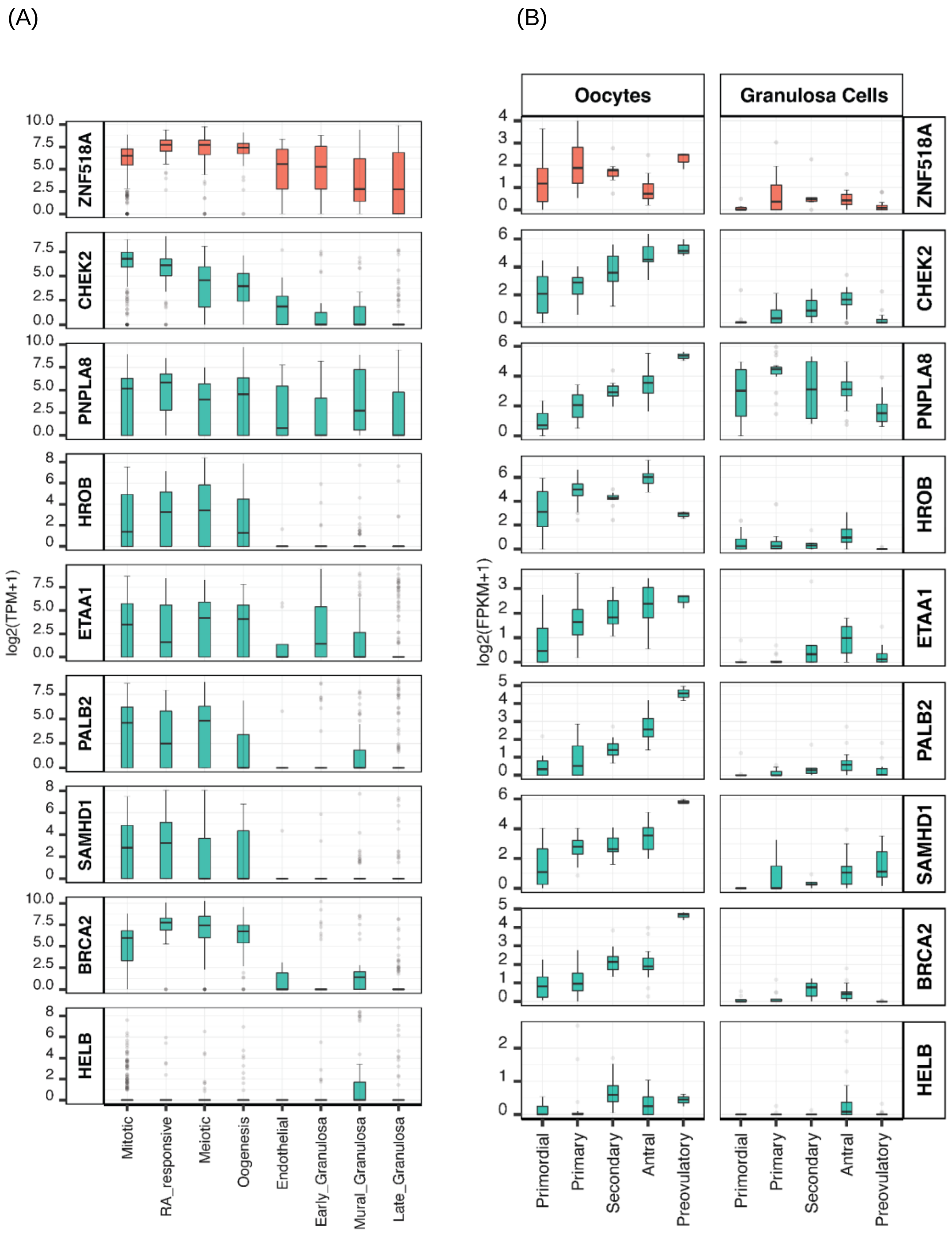


***Supplementary Figure 6: mRNA expression of WES genes during foetal stages and folliculogenesis.*** *Box and whisker plots of mRNA expression of the WES genes at different stages of germ cell development. The plots represent the interquartile range of TPM values, the line at the centre of the box representing the median, error bars indicate the 95% confidence interval and outliers shown as dots.* ***(A)*** *The sub-clusters from single foetal cells from week 5 to 26 post-fertilisation are on the X-axis with the average TPM expression values log2(TPM+1) on the Y-axis.* ***(B)*** *Different stages of folliculogenesis in oocytes and granulosa cells are represented on the X-axis with their average expression values log2(FPKM+1) on the Y-axis.*

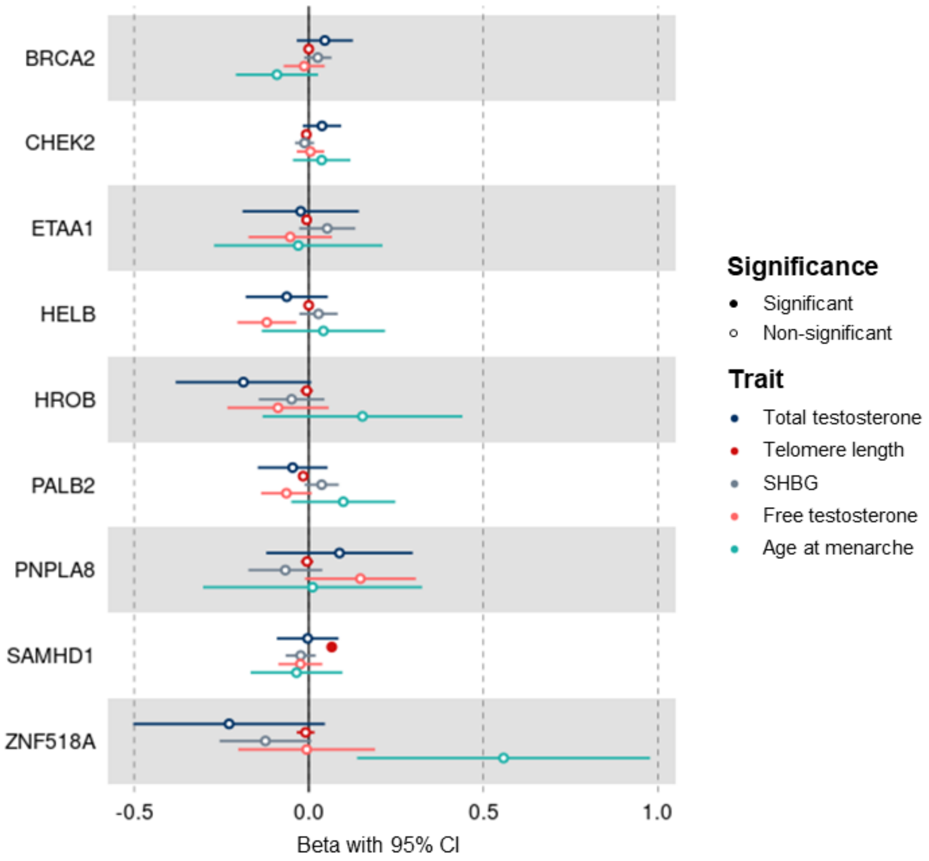


***Supplementary Figure 7: ANM gene burden associations with reproductive ageing-related traits of interest in females only.*** *The coefficients and 95% CIs were female-specific and plotted for the quantitative traits only. The association was tested using BOLT-LMM. Male-specific and sex combined associations could be found in Supplementary Table 10.*

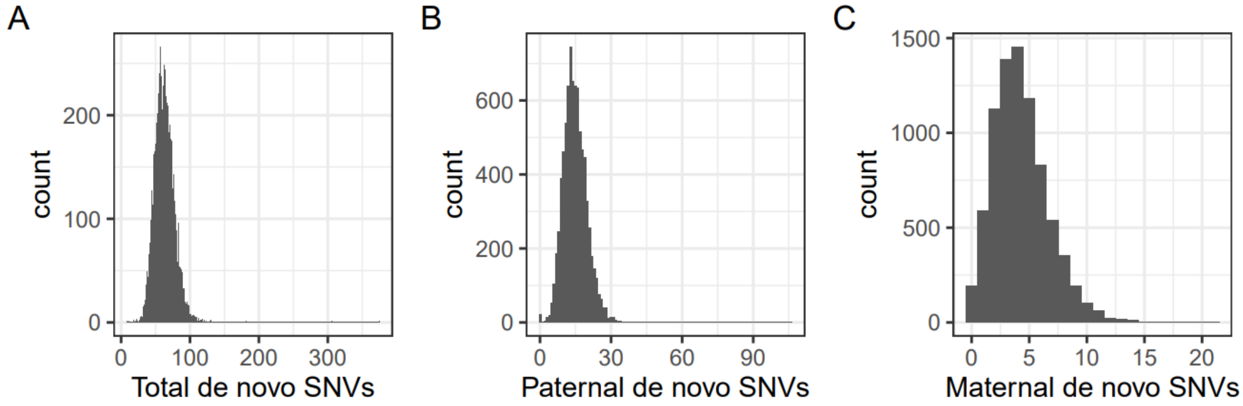


***Supplementary Figure 8: Distribution of de novo single nucleotide variants (dnSNVs).*** *The histogram shows the number of* ***(A)*** *total dnSNVs,* ***(B)*** *paternally derived dnSNVs and* ***(C)*** *maternally derived dnSNVs in unrelated probands with European ancestry from the 100,000 Genomes Project.*
