## supplementary note for "Genetic susceptibility to earlier ovarian ageing increases *de novo* mutation rate in offspring"

### Comparison with previously published results on ANM WES

A previous ANM analysis of the 450K UKBB exome data, published by Ward and colleagues, identified seven genes: *CHEK2*, *DCLRE1A*, *HELB*, *CLPB*, *TOP3A*, *RAD54L* and *HROB*[9]. Our results replicate the association with *CHEK2* and *HELB* and provide more robust evidence ($P$ = $1.9*10^{-8}$) for the previously described suggestive *HROB* association ($P$ = $2.9*10^{-6}$). In addition, we identified six genes, which were not captured by Ward *et al*.: *ZNF518A*, *BRCA2*, *ETAA1*, *PALB2*, *PNPLA8* and *SAMHD1* (**Supplementary Table 3**). We investigated potential study design differences that could account for variation in the findings as the same data was used in both studies.

### Associations not captured in current analysis

First we investigated potential analytical parameters that could account for differences in the findings. Associations with *DCLRE1A*, *RAD54L*, *TOP3A* and *CLPB* were not identified in our study, because we restricted our analysis to variants with a minor allele frequency <0.1%, rather than <1%. We re-analysed our data with a burden test MAF threshold of <1% and three of the four associations were replicated: *DCLRE1A* ($P_{\text{MAF 1\%}}$= $3.8*10^{-8}$, N: 1056), *RAD54L* ($P_{\text{MAF 1\%}}$= $6.4*10^{-7}$, N: 1892) and *TOP3A* ($P_{\text{MAF 1\%}}$= $1.5*10^{-7}$, N: 2001). *RAD54L and TOP3A* were genes highlighted by GWAS and the exome association in *TOP3A* was driven by a single, relatively common variant (rs34001746, MAF=0.7%, $P$=$1.63*10^{-10}$). This variant was in linkage disequilibrium with the previously reported lead GWAS SNP (rs569145577, $r^2$=0.92), with little evidence for association after its exclusion ($P$=0.50 for all other missense and PTVs). *CLPB* just missed our *P*-value threshold, but again a single variant (rs150343959, $P$=$8.22*10^{-6}$) was largely driving the association signal - when excluded in leave-one-out analysis, the *CLPB* burden association dropped ($P$=$1.19*10^{-2}$). By including relatively common variants in gene burden masks, single variants can dominate the general functional effect being tested, which could be contributed to by LD with non-exomic functional variants. Therefore in order to be able to make a stronger link between genetic variants and individual genes, we chose to restrict our analysis to rarer variants with MAF <0.1%.

### Associations not captured by Ward *et al*

Differences in MAF thresholds did not explain why our study identified an additional six genes (*BRCA2*, *ETAA1*, *PALB2*, *PNPLA8*, *SAMHD1* and *ZNF518A*) compared with Ward *et al*. We therefore tested differences in the phenotype preparation, tools and variant masks used to test the associations. Four of our six gene burden associations (*BRCA2*, *PALB2*, *PNPLA8*, and *SAMHD1*) were relatively near the borderline of the significance threshold in our analyses in the primary BOLT-LMM pipeline, although *ETAA1* and *PALB2* were just below the threshold in the REGENIE pipeline (**Supplementary Table 3**).

We had a ~20% larger sample size (106,973 post-menopausal women) in comparison to Ward *et al*. (78,311 unrelated post-menopausal women), which would have resulted in more statistical power in our analyses (**Supplementary Table 3**). This was particularly important for *BRCA2*, *ETAA1* and *PALB2* - Ward *et al.* included 63 (19.5%) fewer *BRCA2* and 46 (21.7%) fewer

*PALB2* carriers of rare damaging variants (Ward *et al.* REGENIE analysis vs. our main analysis) and identified the ANM association at these genes only at the borderline of exome-wide significance, $P$=1.55*$10^{-6}$ and $P$=7.47*$10^{-5}$, respectively. Similarly for *SAMHD1*, Ward *et al.* captured 57 (24.3%) fewer carriers in their linear regression model compared with our main analysis, which resulted in association $P$ values of 6.38*$10^{-4}$ in the linear regression model and $P$=8.02*$10^{-6}$ in the time to event analysis. Ward *et al.* used only unrelated individuals in their primary analyses, where we used linear mixed models and were therefore able to include an additional ~19,000 related individuals. Secondly, Ward *et al.* excluded ~2,300 women with ANM <40 and >60 years, while we used the full natural menopause distribution. Finally, the difference in sample size was partly due to differences in phenotype preparation (resulting in an additional ~7,400 women); specifically, we took into account four instances where questions regarding ANM were asked, whereas Ward *et al.* used data from the baseline visit in their main analysis.

As sensitivity analyses and to better replicate the methods of Ward *et al.*, for four of the genes (*CHEK2* and *DCLRE1A* found in both analyses, and the novel associations in *ZNF518A* and *PNPLA8*) we compared results from linear regression in unrelated individuals using MAF<1% with those from a truncated menopause distribution (ANM 40-60 years) and a time-to-event Cox proportional hazards model (**Supplementary Table 3**). For the truncated distribution, all four genes passed the threshold of exome-wide association and, for all with the exception of *ZNF518A*, the association $P$ value was larger (*CHEK2*: 3.3*$10^{-35}$, *DCLRE1A*: 1.3*$10^{-7}$, *ZNF518A*: 8.4*$10^{-11}$, *PNPLA8*: 3*$10^{-8}$) than for analyses based on the full range of ANM. Association statistics from the Cox model (*CHEK2*: 2.4*$10^{-39}$, *DCLRE1A*: 6*$10^{-8}$, *ZNF518A*: 1.4*$10^{-9}$, *PNPLA8*: 5.7*$10^{-10}$) were comparable to those from linear regression models based on the full range of ANM (*CHEK2*: 3.1*$10^{-46}$, *DCLRE1A*: 2.5*$10^{-8}$, *ZNF518A*: 1.2*$10^{-9}$, *PNPLA8*: 1.9*$10^{-9}$).

Finally, differences in variant annotation may also explain some inconsistencies between studies. *ZNF518A* was not reported by Ward *et al.*, which may be because all variants are in the last and only coding exon of the gene, and in some annotations such variants would inappropriately be excluded from being considered as loss of function. We note that another single coding exon gene (*NFIL3*) was not included in the Ward *et al.* publication, but was in our analysis.

Detailed comparisons between our study and Ward *et al.* are available in **Supplementary Table 3**.
